# Using Geographically Weighted Linear Regression for County-Level Breast Cancer Modeling in the United States

**DOI:** 10.1101/2022.03.28.22272969

**Authors:** Srikanta Banerjee, Matt Jones

## Abstract

Due to the continued disparities in breast cancer, improved models are being needed to inform policy related to existing social disparities related to cancer. First ordinal least squares regression was used to determine the relationship of sociodemographic measures (i.e. poverty rate and social inequity) on breast cancer incidence in the United States. Gini coefficient was used as a measure of income inequality. Next, Geographically Weighted Regression (GWR), a local spatial model, was used to explore the impact location has on the relationship between sociodemographic measures and breast cancer. Mappings of the results are presented, which can assist policymakers to address inequities and social determinants when funding cancer interventions. The GWR model is then compared to linear regression models that do not take into consideration location, highlighting the benefits of spatial models in cancer policy research. More studies applying spatial regression techniques are needed in order to accurately inform policy.

## 1. Introduction

Statistical models to understand health inequities has been of renewed interest in recent years. More specifically, due to increasing concerns in the area of public health, appropriate resource allocations according to health disparities in specific geographic regions is of particular concern. While cancer mortality is declining, there are certain geographic hotspots of age-adjusted cancer-related deaths within the United States. Overall, cancer related deaths continue to be high, second only to cardiovascular disease.

Breast cancer is a common type of cancer among women. More specifically, social disparities continue to exist with breast cancer. Lack of physician availability can lead to lower screening rates in areas where there are individuals who are part of lower socioeconomic status. However, other researchers have attributed socioeconomic deficit to lack of social support, low education, legacy of discrimination, and racial segregation (Braveman & Gottlieb, 2014). These factors have shown to be associated with socioeconomic disparities through psychobiologic pathways. Geographic disparities in breast cancer also exist and is illustrated in Figure 1. Data over geographical regions and time have gained considerable interest in recent years due to increasing concerns of public health, health disparities, and scarce health resource allocations. Ethnic disparities have been documented as well. For instance, it has been found that Non-Hispanic Blacks are diagnosed with triple negative breast cancer more often than other ethnic or racial groups in the United States (Bartley et al., 2020)

**Figure 1:**
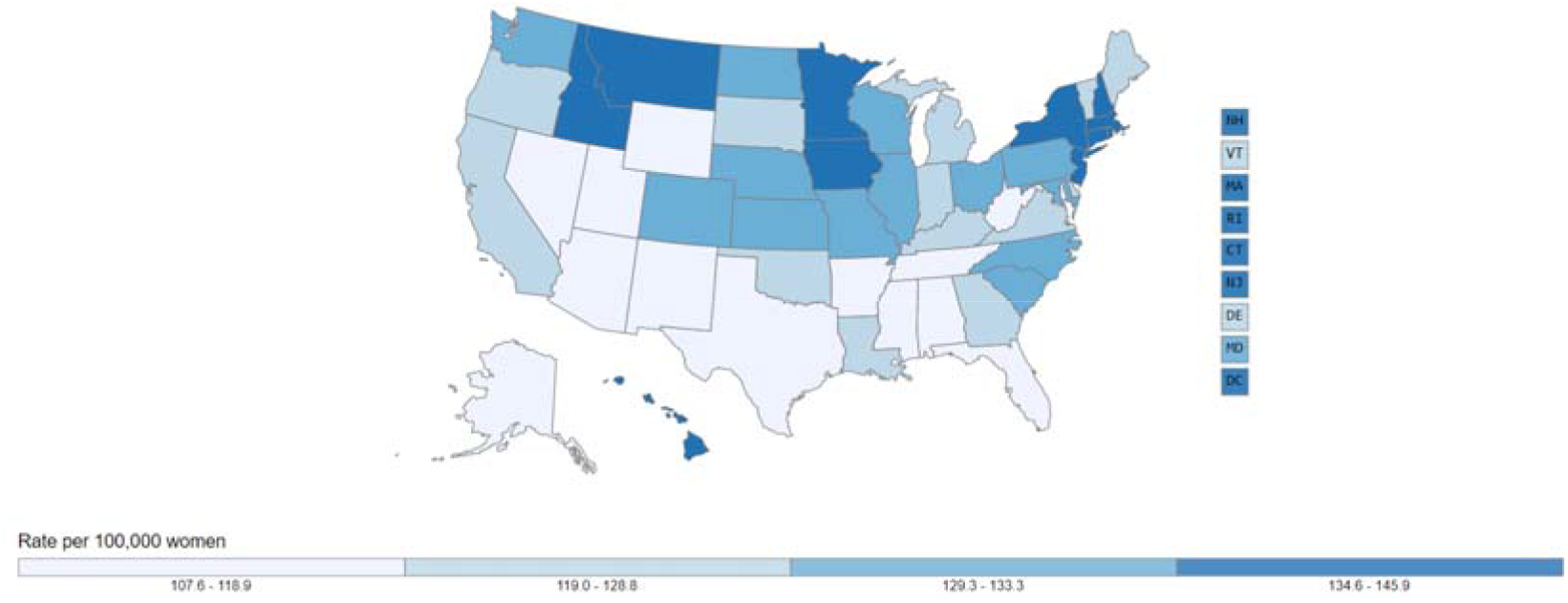
New cases of breast cancer in 2017 in the United States (per 100,000) (CDC, 2020)

The United States Cancer Statistics (USCS) is a comprehensive surveillance system that combines CDC’s National Program of Cancer Registries (NPCR) and the National Cancer Institute’s (NCI) Surveillance, Epidemiology, and End Results (SEER) Program, as well as mortality data from CDC’s National Center for Health Statistics (Bartley et al., 2020; Sharma, 2020). Among many things, the USCS publishes time trends of age-adjusted cancer death rates for different cancer types, and for different sub-populations defined by geographic and socio-demographic characteristics. However, the impact of cancer surveillance is not uniformly effective over geographical regions; see, for example, Figure 1 displays state-level breast cancer incidence rates and the overall distribution for the nation. One of the scientific objectives of monitoring breast cancer rates is to detect changes in the trend over time and identify clusters of sub-populations (generally a set of geographical sub-regions) that are affected by changes (increase or decrease) in risk. Also, if covariates have changes in trend over time, it is interesting to understand how those covariates affect the cancer rates over time through building robust spatially lagged models. A carefully developed procedure that addresses these issues can help administrators find key information for the prevention of breast cancer.

The paper is organized as follows. In Section 2, we describe and compare the concepts of spatial autocorrelation and spatial non-stationarity. Section 3 outlines the sources of data useful in geospatial analysis. Section 4 describes how Moran’s I is informative as a measure of spatial autocorrelation for breast cancer incidence Section 5 describes the statistical properties of global models such as OLS and Poisson Regression. Section 6 provides a robust comparison between OLS regression and GWR. We conclude in Section 7 with interpretation of our results.

## 2. Concepts of Spatial Autocorrelation versus Spatial Non-Stationarity

The concepts of spatial autocorrelation and non-stationarity are critical to understanding how geography affects an individual measure. Spatial autocorrelation is a measure of correlation or similarity between nearby observations. The level of spatial autocorrelation describes the degree to which values at spatial locations (whether they are points, areas, or raster cells), are similar to each other. These locations can be points, areas, or even raster cells.

In measuring something over space, for example the household income, it is likely that two observations that are close to each other in space are also similar in measurement. This assumption is also known as the First Law of Geography: “everything is related to everything else, but near things are more related than distant things” (Tobler, 1970; Albery et al., 2020). This analysis will be completed using Moran’s I measure.

Distinct from spatial autocorrelation is the notion of spatial non-stationarity. Spatial non-stationarity is a condition in which a simple “global” model cannot explain the relationships between some sets of variables. The nature of the model must alter over space to reflect the structure within the data (Brundson et al., 1996; Mollalo et al., 2020). The relationship being modeled is non-stationary throughout space. The Geographically weighted regression (GWR) is a technique mainly intended to indicate where non-stationarity is taking place on the map, that is where locally weighted regression coefficients move away from their global values.

## 3. Sources of Data for studying spatial autocorrelation and spatially lagged models

In conducting the analysis, optimal granularity of the spatial data must be determined. Breast cancer county level age-adjusted incidence rate data from 2015 was derived from SEER*Stat. Typically, the level of granularity is determined by the robustness of the surveillance data available. The poverty rate is defined as the percentage of the population in a county below the county poverty level in the year 2010, a threshold that varies by size and age composition of the household ($25,110 for a four-person household in 2010). The poverty rate was ascertained at county level from Census data. Additionally, Gini coefficient data was applied at the state level to construct the GWR model.

## 4. Spatial Autocorrelation and Spatial Clustering

Using specific statistical measures, spatial dependent can be established. In this model, spatial autocorrelation was tested to determine if breast cancer rates located geographically close to each other were more similar than those rates located far away (Moran’s I = .01, z-score = 36.6, p <. 001) as illustrated in Figure 2. A spatial correlogram of spatial lags was constructed by plotting Moran’s I values as it relates to increasing spatial lag is demonstrated in Figure 3. The spatial lag is a variable that averages the neighboring values of a location. As you can see, spatial autocorrelation is taking place, from the Moran’s I value demonstrating clustering. As expected, as the lags increase, the Moran’s I value starts to decrease since values spatially distant are more dissimilar. In comparison with variograms, another statistic used in spatial statistics, the correlogram from Figure 3 is very useful as an exploratory and descriptive tool. For our purposes, the correlogram plot actually provides richer information than variograms.

**Figure 2.**
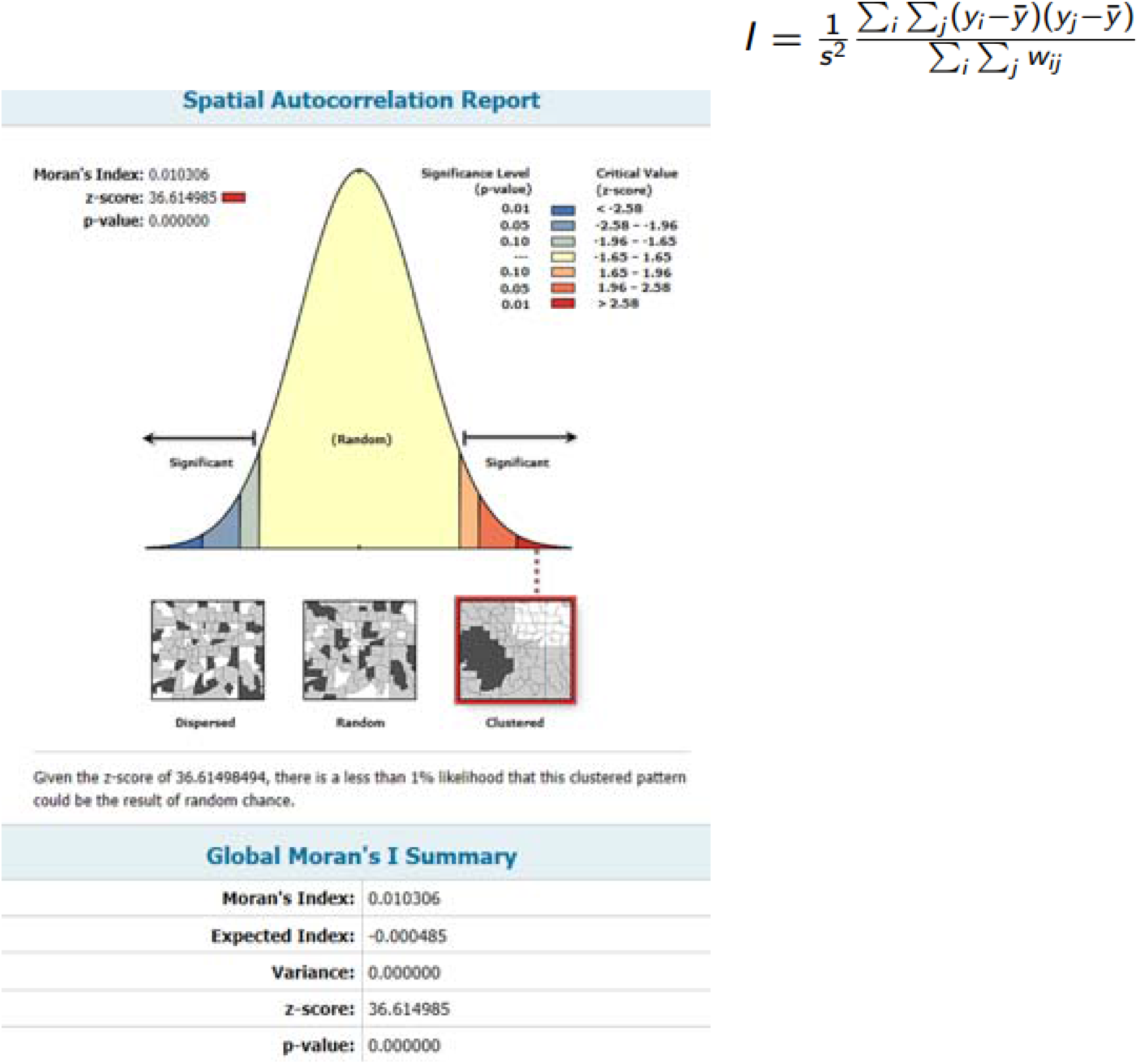
Moran’s I equation and summary statistics for breast cancer incidence variable.

**Figure 3:**
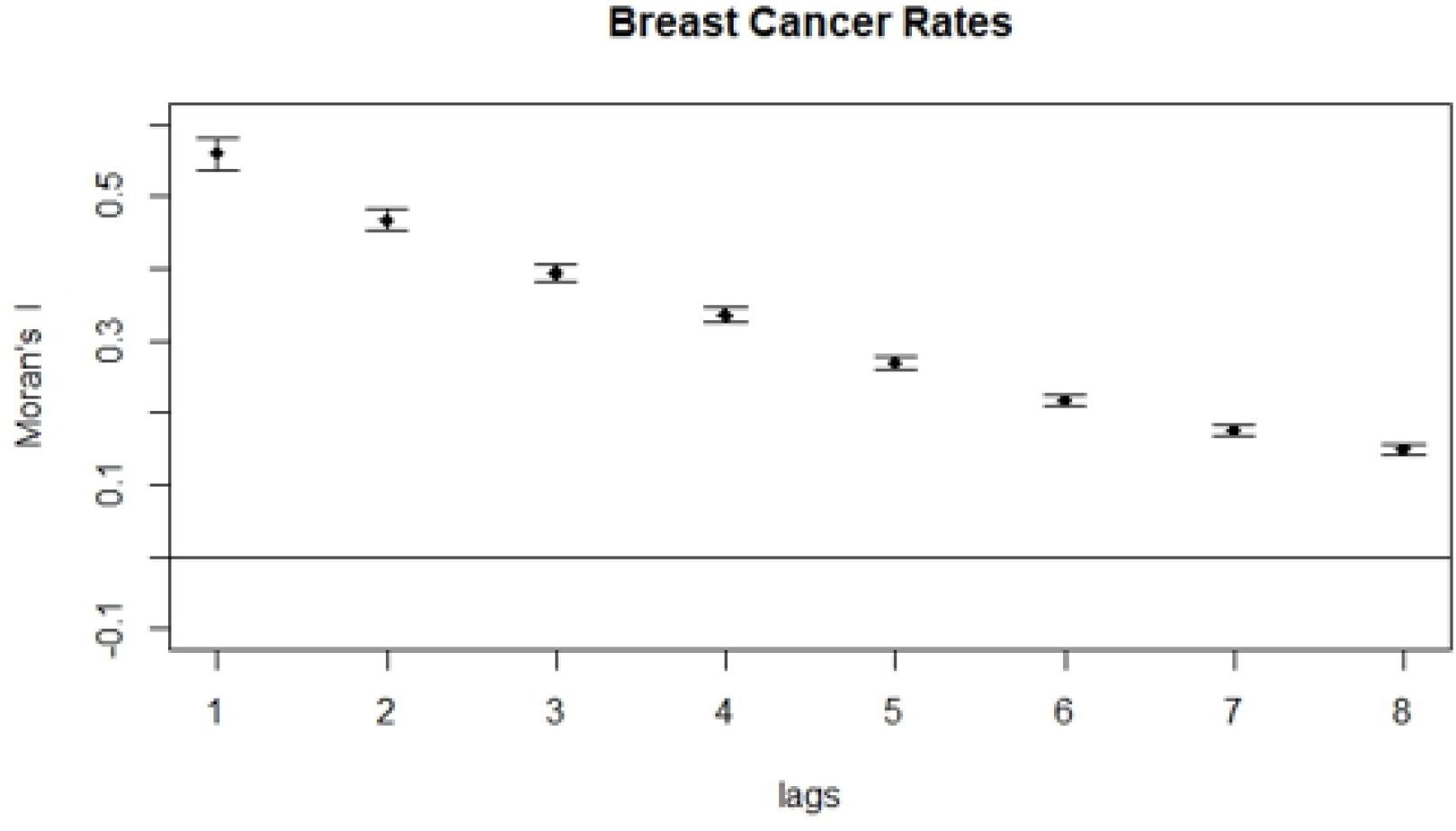
Spatial Correlogram of breast cancer incidence rates with increasing lags.

In order to assess if there are hotspots in certain areas, a Getis*Ord Gi analysis was conducted. The clustering of high breast cancer incidence can be easily seen in Figure 4. A high z-score and small p-value for a feature indicate a spatial clustering of high values. This can be seen in the Northeast and Northwest. A low negative z-score and small p-value indicate a spatial clustering of low values. The higher (or lower) the z-score, the more intense the clustering. The hotspots indicate the need to run spatially lagged variables.

**Figure 4:**
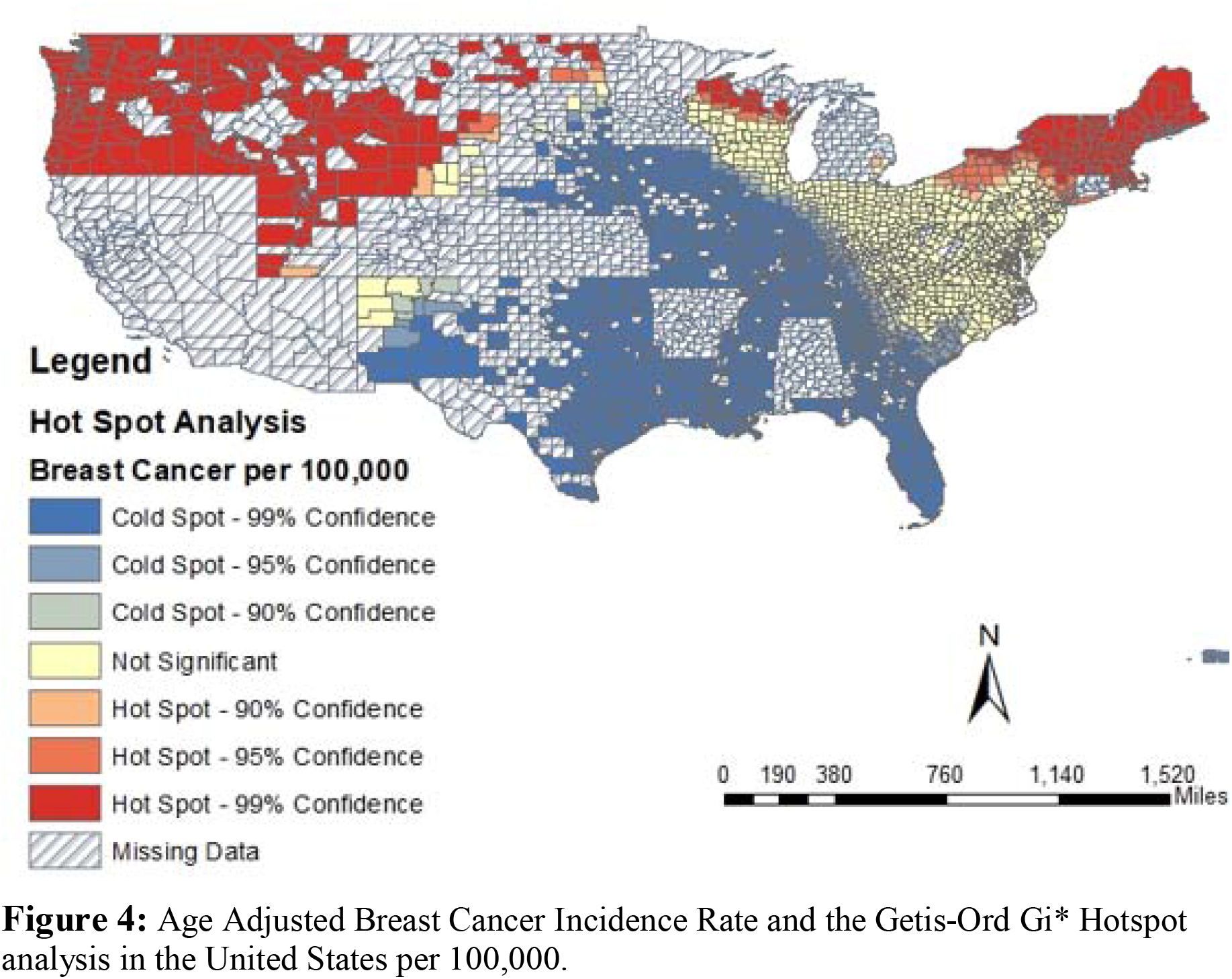
Age Adjusted Breast Cancer Incidence Rate and the Getis-Ord Gi* Hotspot analysis in the United States per 100,000.

**Figure 5:**
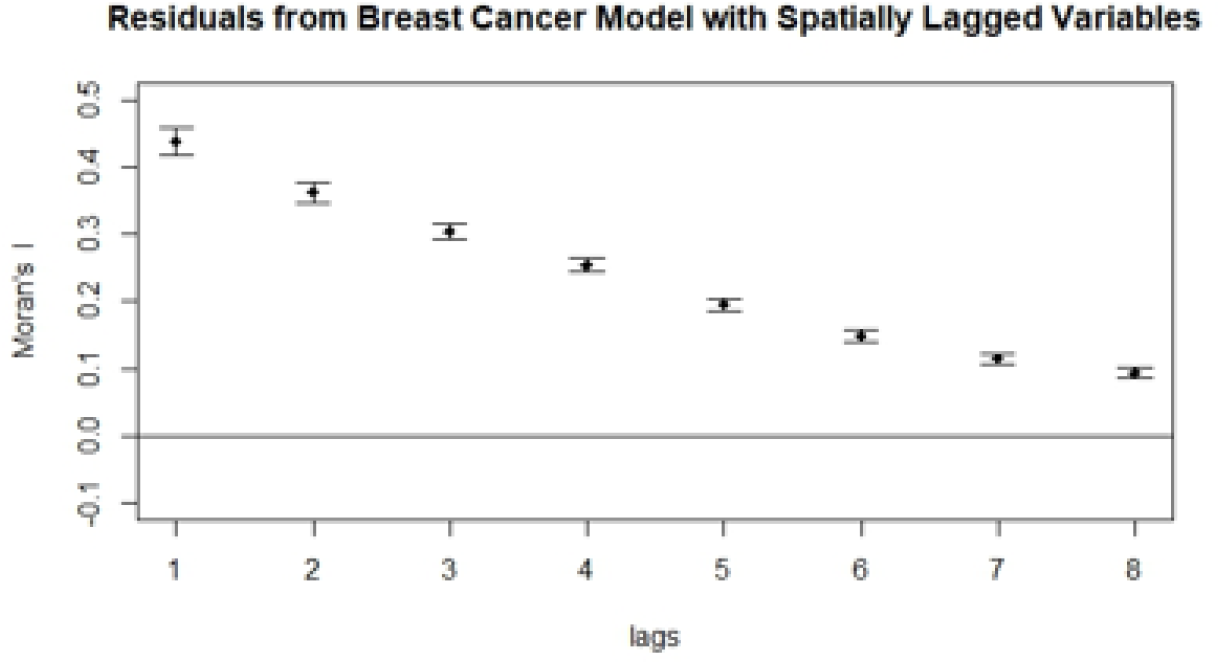
Spatial Correlogram of Residual from Breast Cancer Model with Spatially Lagged Variables.

## 5. Global Regression Models

OLS regression and other regression model that does not account for spatial variation will be referred to as global models. In this paper, we ran some of the more common regression models in order to establish how the relationship between poverty rates and Gini coefficient as it relates to breast cancer incidence rates. Along with OLS regression, Poisson regression is another model appropriate for rate data.

## 6. OLS Regression versus Spatially-Lagged Regression Models

The standard global linear regression model is a global model that does not account for spatial variation and produces a single coefficient of determination (R^2^) and a single β coefficient for each covariate. By contrast, GWR models the relationship between dependent and independent covariates as a series of local models. Each observation is modelled separately, and only includes other observations within a neighbourhood (kernel) based on a fixed distance, or an adaptive distance (based on sample point density) from the modelled observation where points closer to the regression point are weighted heavier than those further away. A GWR model produces multiple coefficients of variation and β coefficients, one for each observation in the sample, and can be expressed formally as:

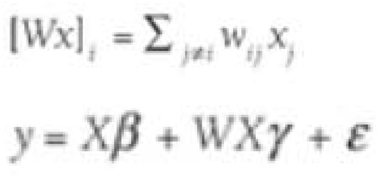

Therefore, the (1) OLS equation and (2) GWR model that is assessed is as follows:

1. Age Adjusted Breast Cancer Incidence Rate per 100,000 = β_0_ + β_1_*Gini Coefficient + β_2_ * Poverty Rate * + Error
2. Age Adjusted Breast Cancer Incidence Rate per 100,000 = β_0_ + β_1_*Gini Coefficient + β_2_ * Poverty Rate + β_3_*Spatially Lagged Gini Coefficient + β_3_*Spatially Lagged Poverty Rate + Error

While many spatial software assume that (by default) spatially lagged dependent variables, as has been pointed out by previous researchers, spatially lagged independent variables can provide further insights about geographical weighting when understanding policy influences. As can be seen in Table 2 of model coefficients, the spatially lagged Gini Coefficient variable and poverty rate variable are both statistically significant. However, more importantly, when the R^2^ between OLS regression (R^2^=0.17) and geographically weighted regression (R^2^=0.23), the geographically weighted model performs more superiorly than OLS regression.

**Table 1:** OLS Regression (R^2^=0.17) and Poisson Regression relating Gini Coefficient and poverty rate to breast cancer incidence data.

|  | OLS Regression |  |  | Poisson Regression |  |  |
| --- | --- | --- | --- | --- | --- | --- |
| Coefficients | $\beta$ Estimate | SE | p-value | $\beta$ Estimate | SE | p-value |
| Intercept | 141.9 | 7.42 | <0.001 | 5.09 | 0.003 | <0.001 |
| Gini Coefficient | 361.4 | 17.61 | <0.001 | 1.18 | 0.005 | <0.001 |
| Poverty Rate | 6.36 | 0.43 | <0.001 | 0.02 | 0.0001 | <0.001 |

**Table 2:** Local Model Coefficients for GWR Model (R^2^=0.23).

| Coefficients | $\beta$<br>Estimate | SE | t-value | p-value |
| --- | --- | --- | --- | --- |
| Intercept | 91.83 | 8.14 | 11.28 | <0.001 |
| Gini Coefficient | 209.69 | 19.83 | 10.57 | <0.001 |
| Poverty Rate | 4.72 | 0.55 | 8.51 | <0.001 |
| Spatially Lagged Gini Coefficient | 0.48 | 0.10 | 4.71 | <0.001 |
| Spatially Lagged Poverty Rate | 80.02 | 5.44 | 14.72 | <0.001 |

## 7. Conclusion

Gini coefficient and poverty rates significantly predicted age-adjusted breast cancer incidence rates per 100,000 in the OLS regression model. The results of OLS regression indicated the two predictors explained 17% of the variance. However, the results of the geographically weighted regression indicated the four predictors explained 23% of the variance, clearly making this a better model fit than a global model like OLS regression. Spatial Autocorrelation test, as tested using Moran’s I, established that geographical considerations were necessary in our final model. Geographical weighting is important when considering cancer-related characteristics (i.e. thyroid cancer, colon cancer, lung cancer, and breast cancer).

Breast cancer is a major public health concern and a significant source of morbidity and mortality. The results demonstrate that there is spatial dependence exists for the breast cancer morbidity, poverty rate, and Gini coefficient variables. In OLS regression, Gini coefficient had a weakly positive relationship with breast cancer rate. Counties with high inequity was associated with counties with higher rates of breast cancer. These findings reflect the tenets of the Theory of Fundamental Causes (Kerstetter & Green, 2014; Phelan, Link, & Tehranifar, 2010). More specifically, despite better screening for cancer, problems with access to medical care continue to impede cancer survival and lead to higher rates of incidence in certain socioeconomic groups (Saldana-Ruiz et al., 2013). Even when controlling for poverty rates, higher inequity leads to higher rates of breast cancer.

Not accounting for spatial autocorrelation can result in biased model estimates and the inability to fully illustrate the possible implications of spatial dependence. Incorporating spatial dependence allows for a better understanding of disease prevalence and correlates. In turn, providing policy makers and health educators with a more accurate understanding of their populations and risk factors is of utmost importance. This understanding can contribute to a more targeted and effective approach to mitigate the effects of disease prevalence. From a policy standpoint, spatial models can also help incorporate the effects of inequality that are associated with “place.”

## Data Availability

The dataset used to conduct this novel data analysis is publicly available through SEER and County Health Rankings.

